# Digital transformations in sex work: A scoping review of social media use by female sex workers in Low- and Middle-Income Countries

**DOI:** 10.1101/2025.02.25.25322659

**Authors:** Doreen Sakala, Moses Kumwenda, Blessings M Kapumba, Gracious Jamali, Frances M Cowan, Wezzie S Lora, Nicola Desmond

## Abstract

**Introduction:** Female Sex Workers in Low- and Middle-Income Countries face increased risks of violence, exploitation, stigma, and limited access to healthcare, leading to vulnerability in their communities. Modern communication technologies have transformed FSW interactions with clients, facilitating easier and more accessible connections. This scoping review explores the use of social media by women involved in sex work in Low and middle-income countries to understand how communication technologies have been adopted and exploited to engage with various stakeholders in the industry.

**Methodology:** We conducted a scoping review following a pre-defined eligibility criterion, systematically searching multiple databases (PubMed, Global Health, Medline, CINAHL, Google Scholar) for English articles published between 1991 and 2024. Two reviewers screened all identified articles and eliminated duplicates using Covidence software. Thematic analysis was performed using NVivo 12 to identify key themes.

**Results:** The search yielded 9,442 articles with eight articles meeting the eligibility criteria for inclusion. Social media was reported to provide an opportunity for women to expand their clientele beyond their immediate geographical confines increasing their income. Use of social media reduced women’s exposure to health risks due to reduced physical contact with clients reducing exposure to HIV and other sexually transmitted infections. Social media also increased physical safety through reduced exposure to violence and increased capacity to screen clients prior to any physical contact. Despite these advantages, women who use social media for selling sex face cyber security risks including the non-consensual dissemination of their pictures and videos, online harassment, and bullying.

**Conclusion:** The scoping review found that there is limited research on women’s usage of social media in sex work in LMIC. Social media empowered female sex workers but also posed risks to their autonomy and mental health. Further research is needed to understand the challenges these women face to inform effective policies and practices.

## Introduction

The 21^st^ century has witnessed unprecedented progress in digital communication technologies, outpacing all other technological advancements and reaching nearly 50% in Low and Middle-Income Countries (LMIC) (1). The emergence of new communication technologies has diversified virtual human interface with websites and social media platforms providing opportunities for social interaction (2). Social media platforms have facilitated access to information, connections with others, and the discovery of and engagement with niche communities (3).

The advent of the internet and digital spaces has led to profound impacts on people’s lives, including women involved in sex work. Over time and with the increasing availability of online opportunities, the sex work landscape has transformed in many countries across the globe, from traditional in-person interactions to online environments (4,5). This shift was first observed in the early 1990s when sex workers began to gravitate towards the Internet as a means of selling sex (6). Since then, FSW engage in selling sex both online and offline, with a growing number increasingly transitioning to online platforms (7).

Online sex work falls into two categories: online service delivery such as webcam modelling, phone sex, and virtual reality experiences (8) and, when the websites are used to market sex, screen clients and make appointments, but sex is sold offline and in person with the client (9). FSW who use the first category are exposed to fewer risks than those who use the second category since there is no physical contact with clients. In High-Income Countries (HIC) where selling sex online is common, the associated benefits include a wider clientele and the ability to pre-screen clients before deciding to meet with them (6). Some websites and social media platforms provide clients of sex workers the opportunity to post reviews which determines FSW scores and their prospects for attracting more clients (10). Social media has also connected FSW with their peers for friendship and support, including discussing and sharing health concerns and safety issues (11). For FSW who rely on marketing sex online only, social media interactions offer protection from both physical harm including getting mugged or raped (12) and risks such as sexually transmitted infections (STI), in. Despite the widespread availability of the internet, low-income countries remain underserved in terms of access and usage (13). In 2022, 92% of the population in high-income countries used the internet, compared to only 56% in low-income countries (14). There is limited empirical research data on the use of social media within the context of sex work in LMIC (15). However, sex work that is facilitated, advertised, performed and accessed via various social media platforms is increasingly popular in LMIC (16,17). For the 2025 fiscal year, the World Bank classifies low-income economies as those with a GNI per capita of $1,145 or less and lower-middle-income economies as those with a GNI per capita of between $1,146 and $4,515 (18). We conducted a scoping review with the aim of understanding how social media is used by women engaged in selling sex and how it promotes engagement in sex work for women in LMIC.

## Methodology

### Overview

We used the Scoping Studies Methodological Framework developed by Arksey and O’Malley (19). This involves identifying the research question, identifying relevant studies, selecting studies, charting the data, and collating, summarising, and reporting. The selection of this framework was based on its ability to accommodate diverse methodological designs within an interdisciplinary field.

### Search Strategy

Two reviewers, DS and GJ, conducted a comprehensive literature search using PubMed, Global Health, Medline, CINAHL, and Google Scholar. These were selected to identify literature in public health, behavioural and social sciences. Global Health and Google Scholar are particularly strong for LMIC studies and studies on marginalised populations such as FSW. We searched for publications using the key concepts: ‘female sex work’, ‘women selling sex’, ‘women’, sex work’, ’social media’, ’Low and middle-income countries’, and their synonyms (see Supplementary file 1). A third reviewer, BK, resolved disagreements on paper inclusion.

Following the initial database searches, we imported all papers identified into Covidence software available at https://www.covidence.org/ to facilitate the identification and elimination of duplicates (20). DS and GJ, independently performed an initial screening of titles and abstracts and then conducted a full-text screening, where 21 discrepancies were identified. To resolve these differences, the reviewers referred to the eligibility criteria for the studies and engaged in discussions, with a third reviewer, BK, who made final decisions for inclusion.

### Eligibility criteria

We included studies published in English between 1995 and 2024 to capture developments since the emergence of social media platforms in the late 1990s. The population of interest was women identified as involved in selling sex, specifically within LMIC. We aimed to include peer-reviewed articles with quantitative, qualitative and mixed-methods designs that focused on complex social phenomena such as the use of social media within sex workspace. However, only qualitative studies were identified on the topic in LMIC and included. We excluded systematic and meta-synthesis reviews.

### Charting the extracted data

DS created a data chart in an MS Excel spreadsheet to describe all selected publications by title of article, author and year, country, study aim, study design, population, social media platform explored and, key findings (Table 1).

**Table 1:**
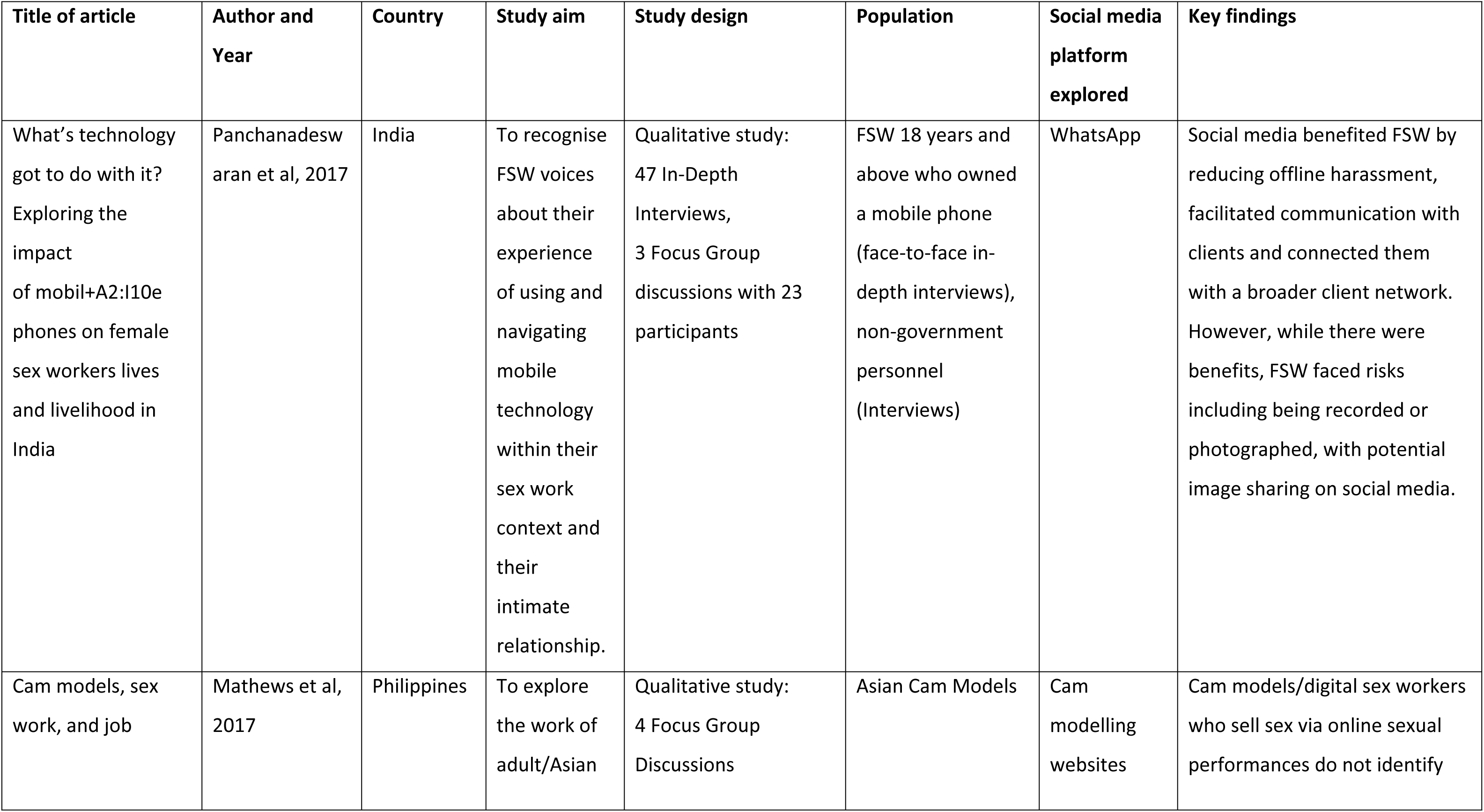
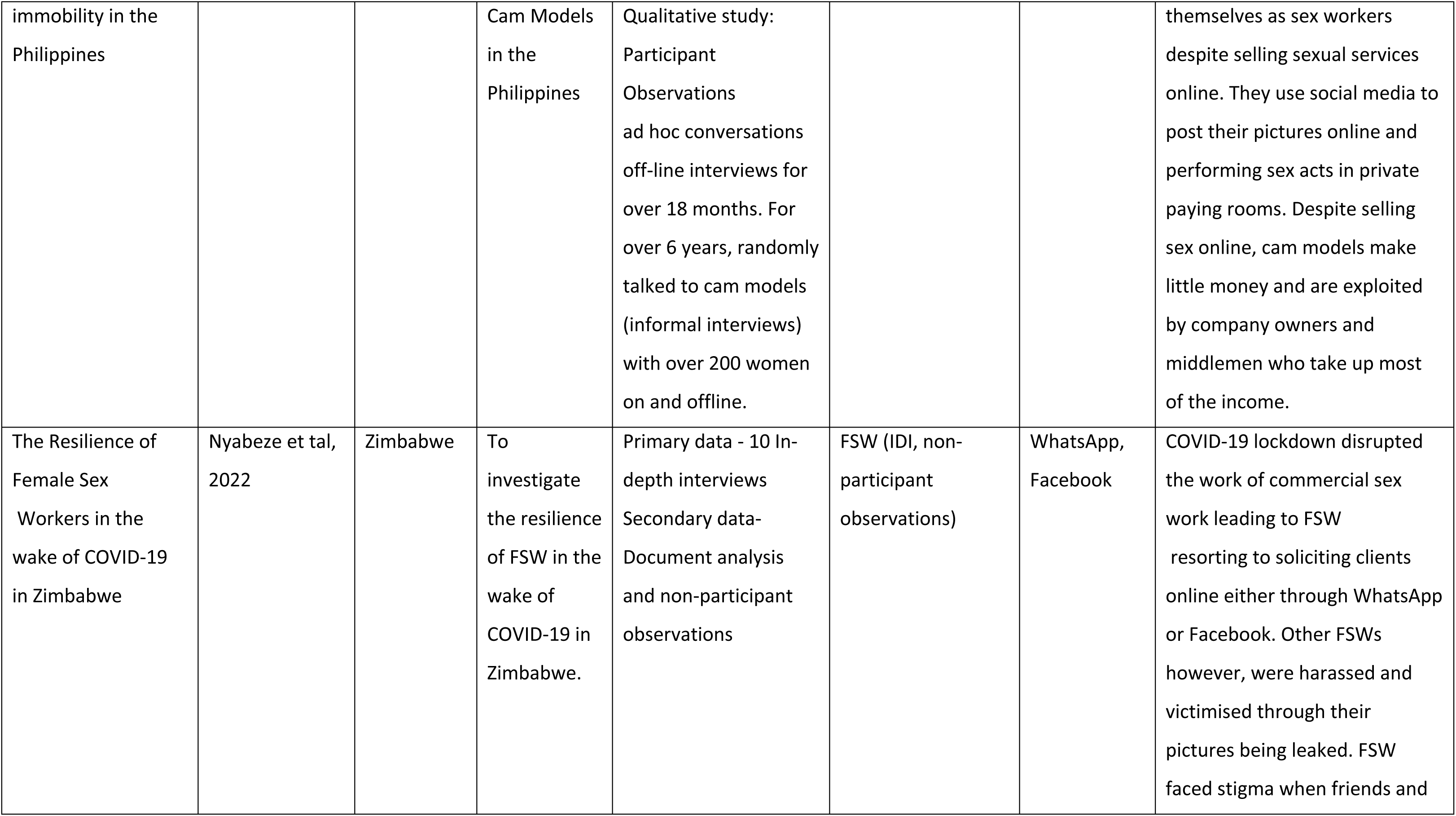
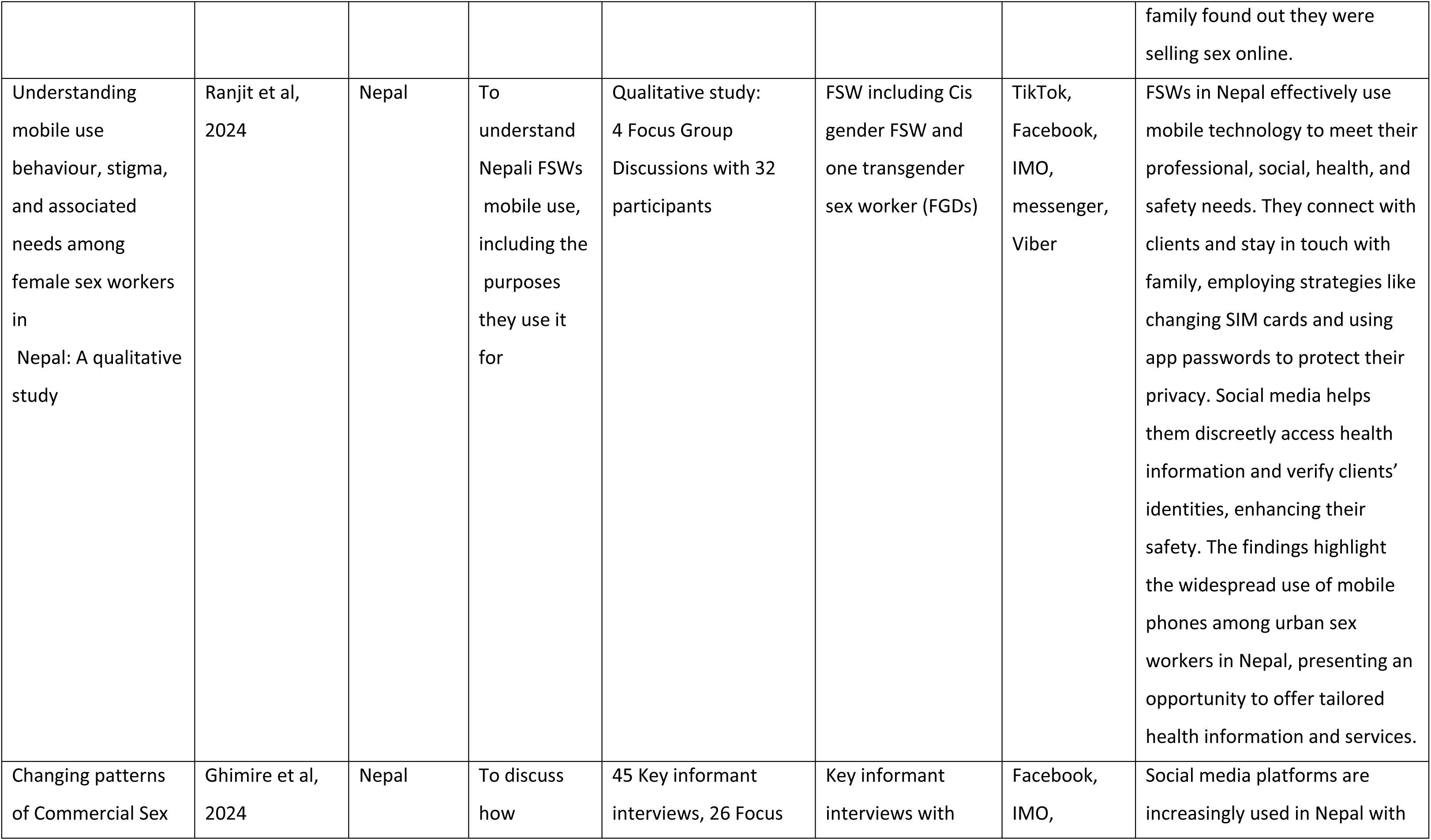
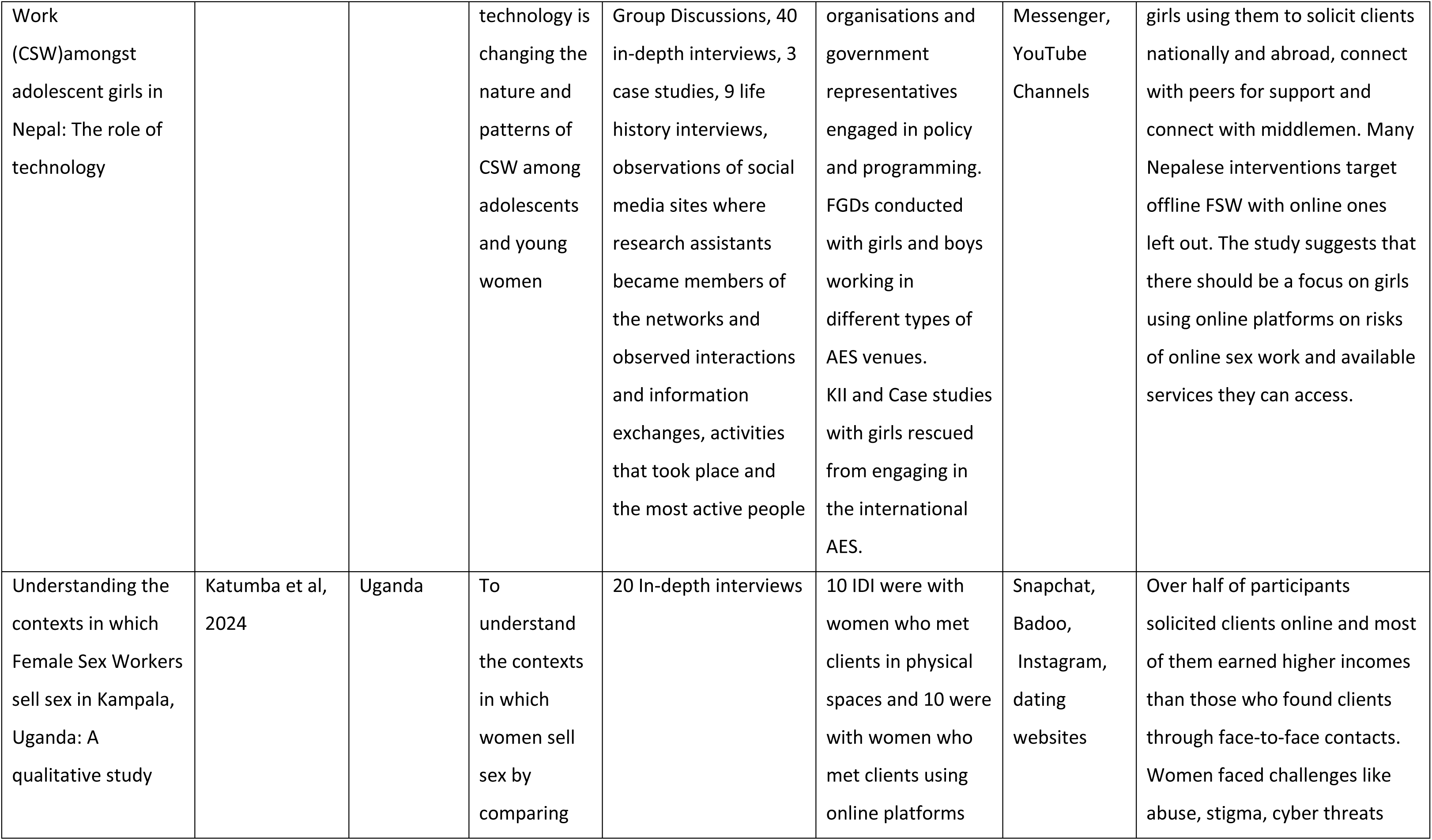
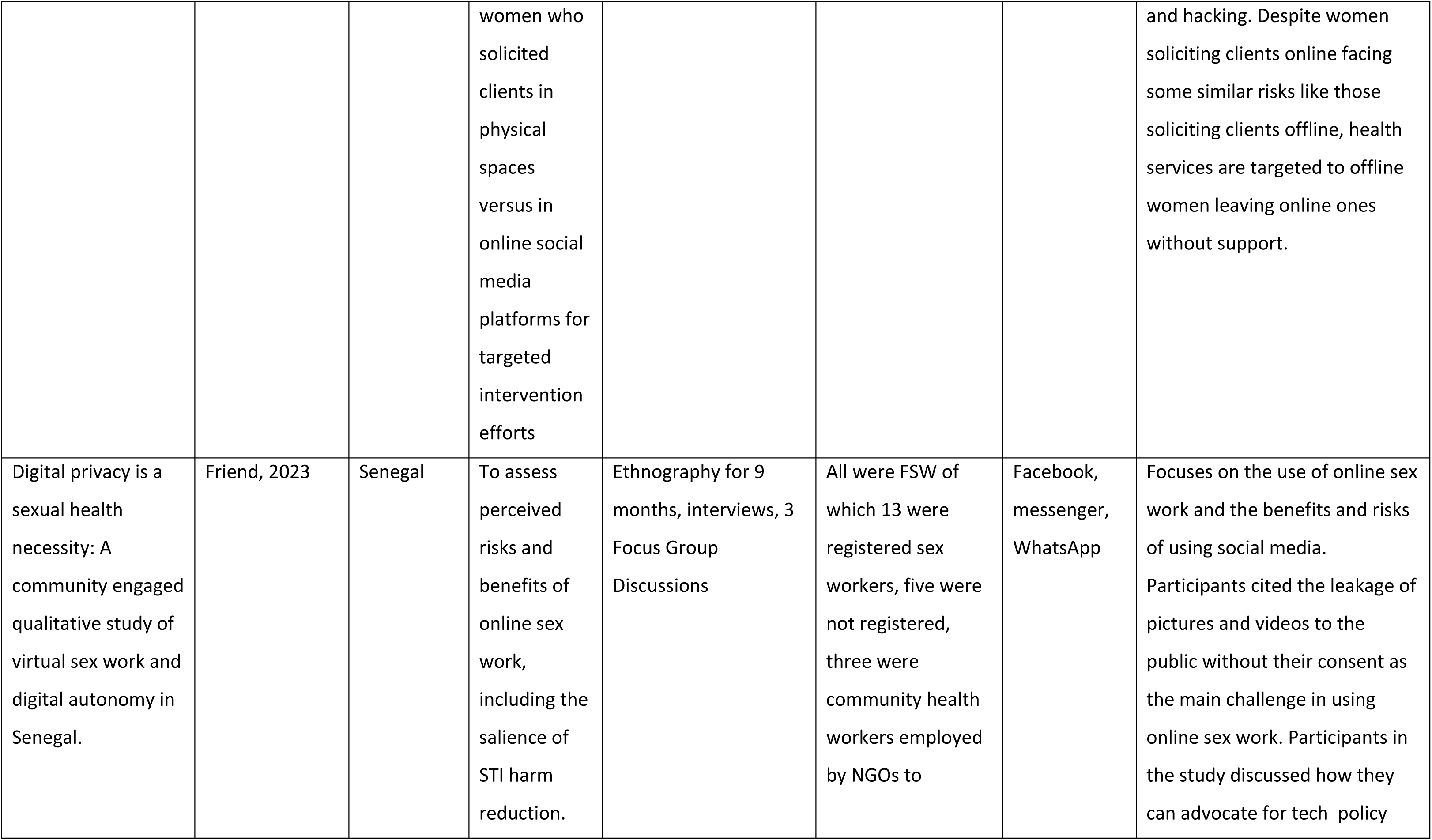
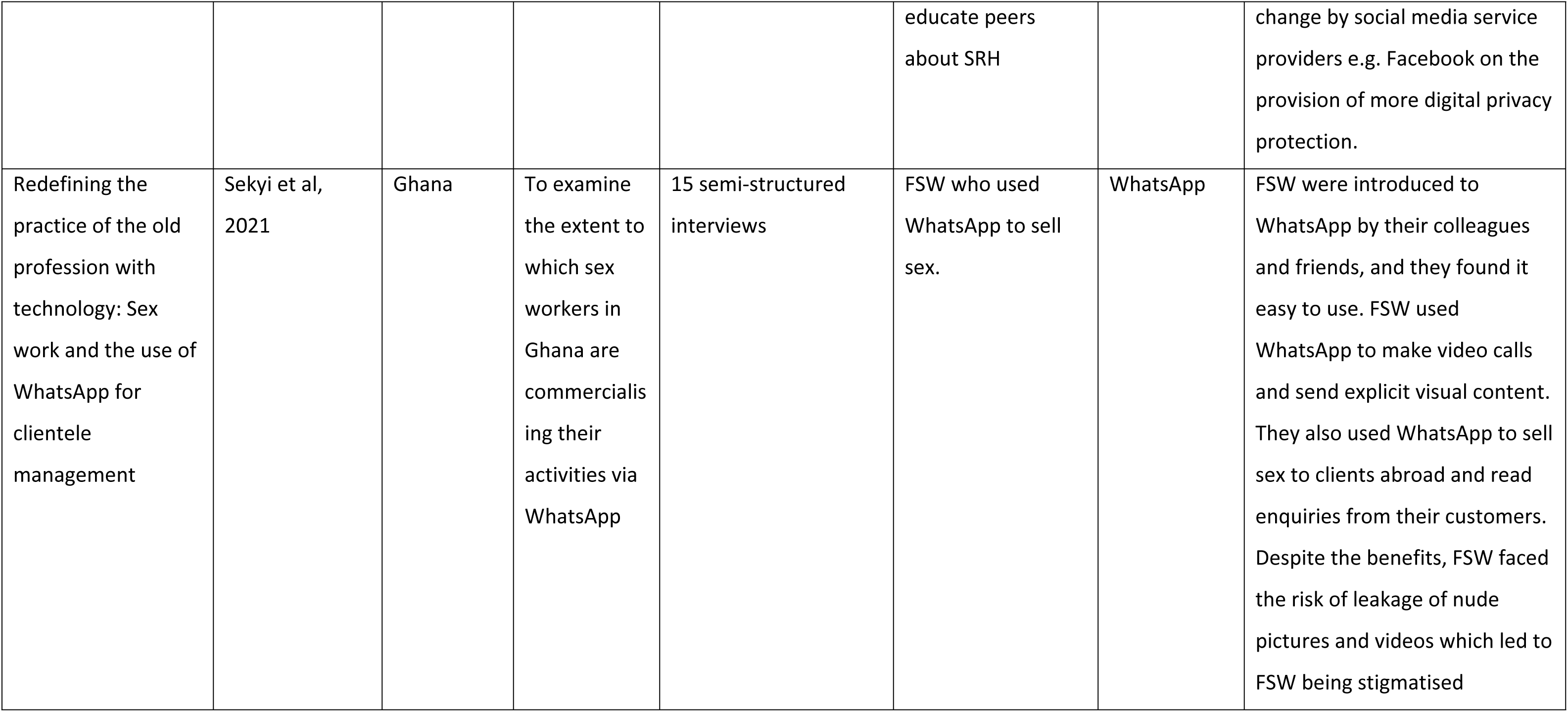
Characteristics of included studies.

### Synthesis of results

The review followed a thematic analysis approach (21), and DS and GJ coded texts line by line from the results, discussions and conclusions of the included studies. We used NVivo 12 (QSR International, Warrington UK), to organise and manage the data extraction process. DS and GJ used an inductive coding approach to identify initial themes. DS and GJ developed second-order themes collaboratively to identify descriptive themes, patterns, similarities, and differences. To ensure accuracy and reliability and trustworthiness, we shared the codes and descriptive themes with all authors for full analysis.

## Results

The search returned a total of 9447 citations, out of which 702 duplicates were identified and removed. The remaining 8745 articles were screened by title and abstract and 8684 articles were removed because they did not report usage of social media for sex work by women living in LMIC but rather focused on a different target population such as Men who have Sex with Men (MSM), adolescents’ usage of social media, the elderly, students, and Intravenous Drug Users (IVDU). We remained with 61 articles for full-text screening and removed a further 53 articles at this stage as they reported outcomes, we were not interested in with others not from LMIC and we remained with eight articles that met the eligibility criteria. Fig 1 details the search and exclusion process and the reasons why some papers did not meet the inclusion criteria. Results were reported thematically with themes aligned with the aim of the study.

**Fig 1:**
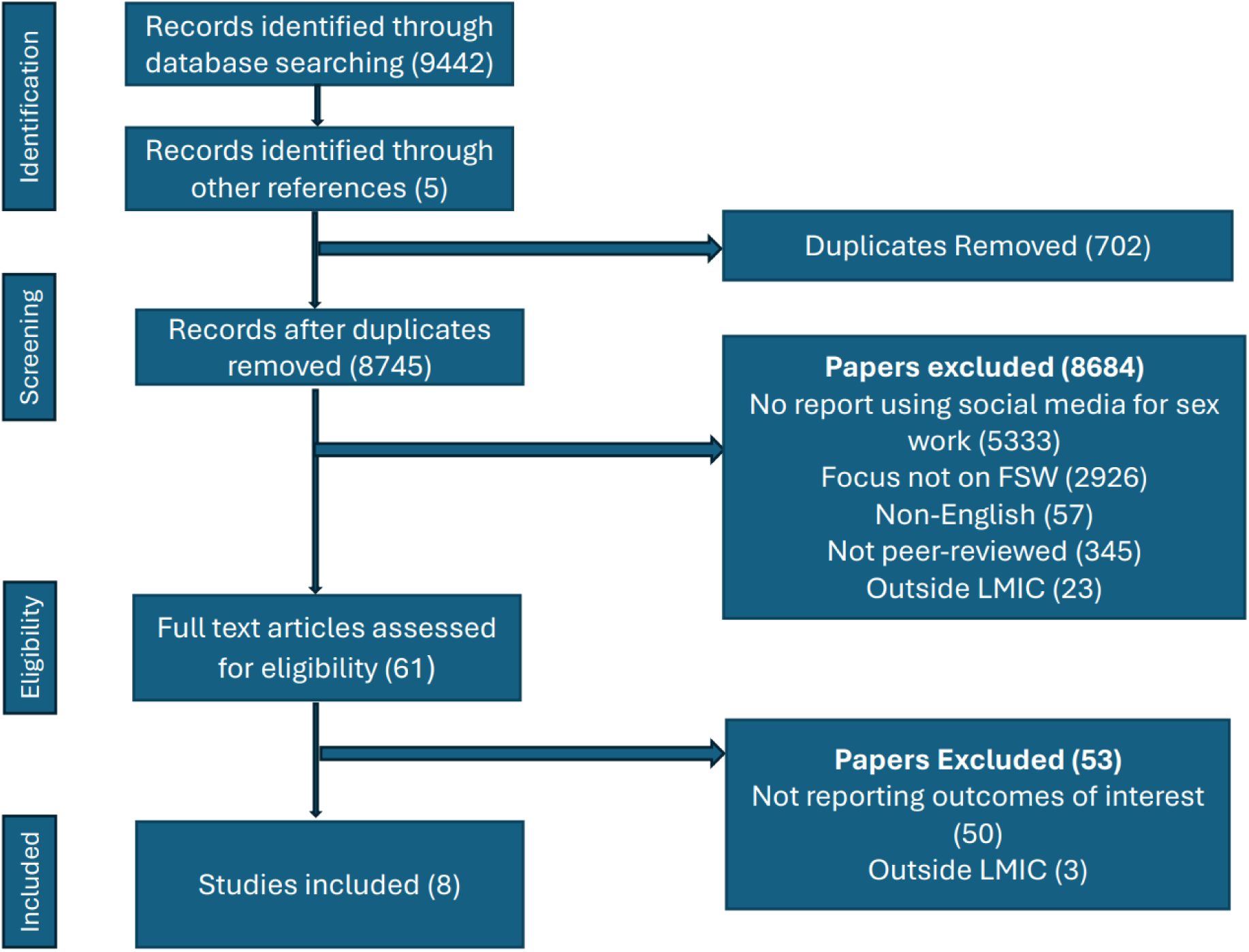
Prisma diagram.

## Results

Four key categories were identified: women’s introduction to online sex work, utility of social media in sex work in LMIC, mechanisms of social media sex work and risks and online sex work. A description of the themes is given below.

### Women’s introduction to online sex work

FSW were introduced to selling sex through social media with friends, colleagues, and clients playing significant roles in this transition (31). For instance, some women learned about online platforms from peers who had already established an online presence, sharing tips on creating profiles, attracting clients, and handling transactions securely (15,23). Clients themselves sometimes suggested moving interactions online for increased discretion and convenience, introducing FSW to specific apps or websites.

During the COVID-19 pandemic, lockdowns, and restrictions such as the early closure of bars and entertainment venues severely limited FSW ability to solicit clients in person. This sudden loss of traditional income sources provoked many FSW to rethink their traditional strategies and quickly adapt in response to the prevailing economic situations by embracing digital communication technologies. In Senegal and Zimbabwe, for example, women began utilising video and audio calls via WhatsApp, an instant messaging application that allows sending of photos, videos, documents, and location. The women also used WhatsApp to offer services remotely, while others turned to Facebook to advertise and connect with potential clients (24,25). These platforms allowed them to reach a wider audience while adhering to social distancing measures. Other FSW developed innovative methods such as online parties or online group chats, to engage clients. However, the shift to online sex work was not without challenges as many FSW faced limitations such as a lack of funds to purchase internet data bundles (23,24). The high cost of data in countries like Zimbabwe made sustained online presence financially burdensome. Poor internet connectivity, especially in rural or underdeveloped areas, hindered consistent communication and could lead to missed opportunities or misunderstandings with clients (24).

### Utility of social media in sex work in LMICs

#### Client solicitation through online platforms

Online platforms that women used for soliciting clients were WhatsApp, Facebook, Snapchat, Instagram, Badoo, dating websites, Facetime, Cam modelling sites, YouTube channels and messenger (15,23–29). Women used the platforms to post their locations, and home addresses, send marketing pictures and videos to clients or use video calls (23–25). Some women who used Facebook, would have conversations with potential clients who would send them a private message and share contact numbers and would continue the conversation on WhatsApp (25). The women would either sell sex online exclusively or arrange via social media platforms to meet the clients in person at a designated location (26) .

The advantage with online sex work was increased income potential when compared to offline sex work (27). Women who engaged in online transactions were often perceived to be of higher social status compared to those who solely conducted in person transactions (26). Among women with lower levels of literacy, voice chats and picture messaging were commonly used, while others were assisted by friends to communicate with clients and schedule appointments on their behalf (15). The quotations below demonstrate how women solicited clients using social media platforms.

> *’As an online strategy, I use WhatsApp to send nude pictures and, at times, pornographic videos to my clients. Clients can also pay us through Ecocash, and in return [we] will pay them in-kind through send[ing] them our nude pictures.* (24)
>
> *“WhatsApp is very good for me because I really like to take pictures and send to my clients…. Some of my men are more interested in my pictures I send to them and the postures I do. In fact some tell me which posture to use in taking the pictures for them and this increases my cash flow.”* (23)

Cam modelling, a recent development in LMIC particularly conducted in the Philippines, involves women going online via a live webcam to perform adult sexual content in front of a person on the Internet and are paid per minute (29). Despite Cam models selling sex online through sexual acts, the Cam models tended not to define themselves as sex workers because of the lack of physical contact with clients. However, street or bar-based FSW felt that cam models are sex workers because they provide sexual services and are paid money out of it (29).

> *For me, being a sex worker is doing it in real, but showing my body in front of the Cam is not being a sex worker.* (29)

#### Extended geographical scope of sex work

Through social media platforms, the exploitation of mobile technology within sex work enabled FSW to expand their client base (27). Before the widespread use of mobile technology, FSW had to physically visit popular solicitation venues such as streets and bars to find clients. However, with the advent of mobile technology, they were able to communicate with potential clients located far away and reach a wider client base in a short time (23,26).

> *‘Previously we had to contact them (clients) on our own, we had to go to bus stand or market to get in touch with customers …. We could not contact customers who were 10–20 miles away from us…. Now it is easy.’* (27)
>
> *“Technology is a good thing that has helped we the ‘sisters’ because it makes everything easy for us. WhatsApp is a good platform to trade for sex and it is even more fun when you get a client who understands the business well. I can do video calls the whole day with some of my clients when they travel out of town”.* (23)

#### Convenience and reduced expenses

A study conducted in the Philippines reported that women who used online platforms to sell sex stated that they saved on transport money as they did not have to move from one place to another (29). Women also stated that online sex work was convenient as they were able to sell sex within their homes, for example, single mothers from Nepal were able to do household chores and take care of their children, and it gave students the flexibility to attend classes (15).

### Mechanisms of social media sex work

#### The use of middlemen

Four out of eight articles described FSW using middlemen to access clients via social media. Middlemen were men or women who acted as a third party to facilitate connections between FSW and clients. The middlemen solicited clients on behalf of FSW using social media platforms or profiles they created, and they comprised of brothel or venue owners, older women who were retired sex workers, the women’s boyfriends and partners, university students who solicited clients on behalf of fellow students, companies as well as pimps and peers (15,26,27,29). The payment was mostly paid directly to the middleman who would take his share or percentage and give the remaining money to the woman. The middleman employed advertising tactics including sending photographs of the women to clients for selection purposes (27).

> *Now, some madams have lots of customers. They click photographs of lots of girls and keep them. The customers insist on viewing the photographs first and then agree on a price. The photographs of the girls are sent via WhatsApp.* (27)

As more FSW became exposed to social media and familiar with its potential, they created their own accounts to independently solicit clients without relying on third parties (27). Women who met clients in person were able to exchange contacts and start engaging on various social media platforms. The women also had the opportunity to receive full payment or negotiate for higher pay as the middlemen were not involved (15). With the use of middlemen, other FSW did not know the amount the client offered to the middleman as they were only given a percentage of it (27).

Some middlemen were involved in blackmailing the women, threatening that they would leak their pictures and videos should they not meet their demands, for instance, asking for a higher percentage of fees from the women. However, middlemen from Uganda for example were able to shield violent customers who posed a risk to the women (26). The advent of social media therefore enabled women to solicit clients on their own and be in charge of their pictures and of the clients to whom they wanted to sell sex. The quote below demonstrates how women benefitted financially by not using middlemen.

> *When I was in the brothel, I did not know how much the clients paid the brothel madam. She used to give me 150/200 rupees and tell me that, that it is the money the client gave. If they sent me out during the night, I did not know how much the clients paid. When we went out in the night, thinking that it is just one man, there would be two or three, and I would face some problems which are difficult to express. So, some of the clients started telling me to take my phone number we can meet outside. I started contacting more clients, I made a house for myself and started building my life.* (27)

#### Flexibility through social media

Women who engaged in online sex found it advantageous due to its flexibility. The women were able to sell sex online within the confines of their own homes and were still able to get on with household and family responsibilities, saving them cost for transportation while promoting a sense of autonomy and independence (25,29).

#### Relationship between FSW and social media clients

Three studies found that using mobile technology contributed significantly to enhancing the relationships that FSW had with their clients by facilitating regular communication (15,23,27). Participants also highlighted the crucial role of consistent communication in sustaining relationships with clients and ensuring continued patronage (27). FSW found social media platforms like WhatsApp empowering as they were able to directly read important messages regarding appointments from their clients (23). Thus, mobile technology empowered FSW to stay in touch with preferred clients, especially those providing financial support (15).

#### Access to health information

One study from Nepal found that social media was beneficial for accessing sexual health information, particularly on sensitive topics which were difficult to discuss in person. FSW often searched for relevant information on different social media platforms before visiting a medical facility, to obtain a better understanding of a particular disease or condition (28). Sexual activity being a taboo in Nepal, women found it beneficial to have an idea about the health problems they may be facing, as per the quotes below.

> *With a mobile, if I need any information I would search and look for it. Before going to the clinic, we will know a little bit about what is going on with me.* (28)
>
> *Before mobile phones, it was embarrassing to ask about sexual health issues and sexual diseases. Now we can search for information on Google and it is also available on YouTube. Information and diseases related to STIs.* (28)

### Harms and benefits of sex work supported through social media

#### Leakage of nude pictures and videos

Four out of eight studies reported women fearing or experiencing the risk of their pictures or videos being leaked by clients (23–25,27). Clients would covertly record videos or take pictures of the women during online sexual encounters. They would then blackmail the women threatening that they would expose or leak the videos they had recorded if they did not pay back the money they had paid.

> *“He wanted to publish. I cried, cried, cried, me alone. I couldn’t tell anyone”-* (25)
>
> *“I always try to hide my face but some of my clients they tell me they want to see my beautiful face. I like WhatsApp but not very much because of the future consequences if it shall happen.”-* (23)
>
> A participant from Senegal compared the risk of having an STI and being exposed on the internet. She cited that it was better to have an STI as one would be able to go to the hospital for treatment and only the health care worker would know about it (25). Being exposed on the internet meant a lot of people accessing one’s pictures or videos which can never be deleted.
>
> *“STIs, you can go to the midwife, she cures you…the Internet is more painful.”-* (25)

Five out of eight studies also showed that women had the fear of their pictures or videos leaked which would lead to societal shame, depression and, stigma (23–27). Increasing access to social media impacted fears and perceptions of risk-related physical encounters for clients that women met online and agreed to meet in person offline. A study conducted in Senegal reported that to avoid being recorded, women discouraged or disallowed the client from going into a room for a sexual encounter with any gadgets (25). Women also faced cyber harassment and hacking of their social media pages where hackers were able to access their personal information (26). Due to such risks, women who started selling sex on social media during COVID-19 in Zimbabwe returned to offline sex work once social interactions were again possible, citing online sex work risks as the reason (24).

There were also profound levels of social stigma associated with sex work especially when their pictures or videos were leaked (23). This brought shame and isolation from their friends, family and society, as they were seen as having no morals. (25).

> *If any members of your family or friends view intimate images of you online, “ah,“ah, your future is broken.” One risks being perceived as immoral and lacking sutura. This could in turn cause rejection by family. Outing could also jeopardise her career aspiration –* (25)

#### Reduced exposure to health and legal risks

Women who sold sex online were noted to be less exposed to the risks of being arrested and abused by police who primarily targeted women selling sex in streets and venues (27). Women who sold sex online only, faced less physical abuse from clients and police officers as opposed to women who sold sex offline. Women who marketed sex online but later met clients in person also faced chances of physical violence. Five out of eight studies reported that sex sold online only was preferred as it did not expose one to contracting sexually transmitted diseases and HIV as opposed to in-person sex work which was associated with risks like clients refusing to wear condoms, removing condoms and accepting to have unprotected sex for a higher pay (25). During COVID-19 women also cited online sex work as the best way to avoid contracting the disease as it limited in-person contact with clients (24).

> *“Body to body you have the risk of HIV. Calls are better, there’s no disease there…calls don’t tire you out. You don’t have to wear heels, you sit in your house in security…your health is secure.”-* (25)

#### Opportunity to pre-screen clients

Through online conversations women had control over the type of client they wanted to transact with as they had time to investigate the client and assess if they should sell sex to him or not (28). The women used different tactics as a way of pre-screening the client to identify potential for harm including using applications like Facetime to identify clients before scheduling a meet-up for security purposes (28). Clients who were respectful were prioritised as opposed to clients who were rude and aggressive as these had a higher chance of being violent or abusive. The women would also not agree to meet clients in their homes but rather in hotels near their location so that they could easily seek help (26). Women also avoided men who negotiated for a price drop as this was also a sign that they may turn out to abuse them.

> *The advantage of hotels is that you can easily get help in case of any problems, which you can’t get when you are in someone’s home because its already night and some people’s homes are fenced even if you shout no one can help.* – (26)

#### Use of false identities and alternative contact numbers for business

It was common among women to have different sim cards or phones where they used one to communicate with family and friends and the other to communicate with customers (28). The same also applied to social media platforms where women would have multiple accounts, one with their real name for family and friends and others with a fake name which they used to communicate with clients (15,25). This was a way of avoiding being recognised as a sex worker.

#### Use of Peer networks

Women involved in sex work formed networks as a way of supporting each other. To prevent abuse, women who agreed to meet a client in person, would notify their friends of the location that they were meeting their clients. They would also deliberately call their friends in the presence of the client and notify them of their location as a way of alerting the client that should they be abused their friends are aware of where they were (22). Other women relied on clients who have been referred by their friends and if it was a new client, they would spend time chatting with the client online as a way of screening them. If they observed any unusual behaviour, the women would not agree to meet the particular client in-person (15).

## Discussion

This scoping review highlights a lack of empirical evidence on social media use within sex work in LMICs. This gap may be attributed either to a lack of research attention in this area or to the limited number of women engaging in sex work through social media in LMIC settings. We found that despite the lack of evidence, social media plays a significant yet under-explored role in reshaping the sex work landscape, and one that was strengthened as a result of COVID restrictions in LMIC from 2020 to 2021. The findings also underscore the transformative impact of social media platforms on the practices, opportunities, and vulnerabilities faced by FSW.

The advent of social media platforms has enabled FSW to shift from traditional, in-person solicitation to more discrete and far-reaching online practices. Platforms such as WhatsApp and Facebook, have emerged as vital tools for client solicitation, facilitating broader geographic reach and reducing the dependency on physical proximity (23–25). This transition aligns with global trends observed in high-income countries, where technology has revolutionised sex work (3,30–32). Yet, limited access to reliable internet connectivity and affordable data bundles remains a barrier, highlighting persistent digital inequalities that exacerbate existing socio-economic disparities (24,33).

Introduction to social media platforms involved middlemen who often managed transactions on behalf of FSW. While this arrangement has facilitated their work, it had challenges including exploitation (22,28). Eliminating these middlemen through social media meant that FSW were empowered to negotiate directly with clients, retain full payments of the transaction, and exert greater control over their work (16). This economic autonomy represents a critical advantage of independent management of social media accounts linked to sex work, but the attainment of this independence demanded FSW to adopt protective strategies such as using pseudonyms and maintaining multiple social media accounts to allow them to reduce the potential for social harms linked to solo online transactions. These tactics demonstrate the resilience and resourcefulness of FSW, as they manage the dual pressures of economic survival and societal judgment.

Social media platforms have become important conduits for accessing a broader array of important information including health information. FSW frequently use social media to gather insights into sexual health issues before seeking medical attention, reducing barriers to healthcare access in conservative or stigmatising environments (28). While this practice empowers FSW to make informed health decisions, it also underscores the need for accessible information that emphasises the importance of seeking professional medical assistance (34).

Despite the benefits, online sex work introduces new risks. Concerns around digital privacy, including the unauthorised dissemination of images and videos, was pervasive and often resulted in severe psychological and social repercussions, such as isolation and depression (23,25). The pervasive social stigma surrounding sex work exacerbates these challenges, with leaked content leading to social isolation and familial rejection (23). Furthermore, online platforms expose FSW to cyber harassment and hacking, necessitating enhanced digital literacy and security measures to protect their identities and livelihoods (24,25).

The findings reveal a perceived trade-off between physical and social media risks. While online platforms reduce exposure to physical violence and sexually transmitted infections, they introduce psychological and reputational hazards that are often more enduring (25). This contrast reflects the complex realities of navigating sex work in the digital age, where safety and vulnerability coexist in new and challenging ways. The risks of online sex work identified in this study, including online harassment, and blackmail, are consistent with existing literature on the vulnerabilities of social media-based sex work (28).

The findings also demonstrated that peer networks play a pivotal role in fostering safety and resilience among FSW. Through these networks, women share strategies for client screening, negotiate safer meeting arrangements, and provide emotional and practical support (28). This sense of community underscores the collective agency of FSW in mitigating risks and enhancing the benefits of their professional experiences.

### Limitations

The scoping review included only eight studies reflecting the limited body of research on women who use social media for sex work in LMIC. Such scarcity of research underscores the need for further studies to provide a comprehensive understanding on the topic. This small evidence may therefore limit the generalisability of the findings and provide an incomplete understanding of the topic. The studies included in the review also did not cover all LMIC regions comprehensively omitting insights from countries where social media use may be prevalent but not documented.

## Conclusion

This review illustrates how women use social media for sex work and the double-edged nature of social media in sex work within LMICs. Social media for sex work is protective from a health perspective as it empowered women, provided them with access to health information and reduced exposure to STI and HIV. It however also exacerbated social risks such as leakage of pictures and videos that lead to stigma affecting one’s psychological well-being and mental health. More work needs to be done to identify the potential role of social media in Sexual Reproductive and Health (SRH) interventions and risk reduction approaches for FSW in LMIC. Future research should also focus on developing tailored interventions to address these risks, including digital literacy programs and robust online safety measures. Policymakers and stakeholders must also work toward creating inclusive digital environments that safeguard the rights and dignity of FSW while leveraging the transformative potential of technology to enhance their safety and well-being.

## Data Availability

All relevant data are within the manuscript and its Supporting Information files.

https://www.covidence.org/

## Acknowledgements

The author wishes to thank the Adapted Microplanning Eliminating Transmissible HIV in Sex Transactions (AMETHIST) consortium where this study was embedded in and the Liverpool School of Tropical Medicine where she is undertaking her PhD studies.

