## Supplementary file 1 for "Digital transformations in sex work: A scoping review of social media use by female sex workers in Low- and Middle-Income Countries"

**S1 Fig: Search Strategy**

| Key words | Search terms |
|---|---|
| Sex work | “woman” OR “WOMEN” OR "female sex work*" " OR prostitut* OR "transactional sex" OR "sex for money" OR "commercial sex" OR “sex” OR “sell sex” OR “sold sex” OR "trad* sex*" OR "commercial sex" OR “sex for business” OR "girls sell* sex" OR "women sell* sex" OR “high risk women” OR “high risk girl*” OR “FSW” OR “key population*” OR “only fan*” OR “cybersex” OR “mobile wom* |
| Low- and Middle-Income Countries | (afghanistan OR albania or algeria OR "american samoa" OR angola or argentina OR armenia OR armenian OR azerbaijan OR bangladesh OR "republic of belarus" OR belarus OR byelarus OR belorussia OR byelorussian OR belize OR "british honduras" OR benin OR dahomey OR bhutan OR bolivia OR "bosnia and herzegovina" OR bosnia OR herzegovina OR botswana OR bechuanaland OR brazil OR brasil OR bulgaria OR "burkina faso" OR "burkina fasso" OR "upper volta" OR burundi OR urundi OR "cabo verde" OR "cape verde" OR cambodia OR kampuchea OR "khmer republic" OR cameroon OR cameron OR cameroun OR "central african republic" OR "ubangi shari" OR chad OR china OR colombia OR comoros OR "comoro islands" OR "iles comores" OR mayotte OR "democratic republic of the congo" OR "democratic republic congo" OR congo OR zaire OR "costa rica" OR "cote d'ivoire" OR "cote d' ivoire" OR "cote divoire" OR "cote d ivoire" OR "ivory coast" OR cuba OR czechoslovakia OR djibouti OR "french somaliland" OR dominica OR "dominican republic" OR ecuador OR egypt OR "united arab republic" OR "el salvador"OR"equatorial guinea" OR "spanish guinea" OR eritrea OR eswatini OR swaziland OR ethiopia OR fiji OR gabon OR "gabonese republic" OR gambia OR "georgia (republic)" OR georgia OR georgian OR ghana OR "gold coast" OR grenada OR guatemala OR guinea OR"guinea bissau" OR guyana OR "british guiana" OR haiti OR hispaniola OR honduras OR india OR indonesia OR timor OR iran OR iraq OR jamaica OR jordan OR kazakhstan OR kazakh OR kenya OR "democratic people's republic of korea" OR north korea OR kosovo OR kyrgyzstan OR |

| | |
|---|---|
| | kirghizia or kirgizstan OR "kyrgyz republic" OR kirghiz OR laos OR "lao pdr" OR"lao people's democratic republic" OR lebanon OR "lebanese republic" OR lesotho OR basutoland OR liberia OR libya OR "libyan arab jamahiriya" OR "republic of north macedonia" OR macedonia OR madagascar OR "malagasy republic" OR malawi OR nyasaland OR malaysia OR "malay federation" OR "malaya federation" OR maldives OR "indian ocean islands" OR "indian ocean" OR mali OR malta OR micronesia OR "federated states of micronesia" OR kiribati OR "marshall islands" OR tuvalu OR mauritania OR mauritius OR moldova OR moldovian or mongolia or montenegro or morocco or ifni or mozambique OR "portuguese east africa" OR myanmar OR burma OR namibia OR nepal OR "netherlands antilles" OR nicaragua OR niger OR nigeria OR muscat OR pakistan OR panama OR "papua new guinea" OR paraguay OR peru OR philippines OR philipines OR phillipines OR phillippines OR rwanda OR ruanda OR samoa OR "pacific islands" OR polynesia OR "samoan islands" OR "navigator island" OR "navigator islands" OR "sao tome and principe" OR senegal OR serbia OR "sierra leone" OR slovakia OR melanesia OR "solomon island" OR "solomon islands" OR "norfolk island" OR "norfolk islands" OR somalia OR "south africa" OR "south sudan" OR "sri lanka" OR ceylon OR "saint lucia" OR "st. lucia" OR "saint vincent and the grenadines" OR "saint vincent" OR "st. vincent" OR grenadines OR sudan OR suriname OR surinam OR "dutch guiana" OR "netherlands guiana" OR syria OR "syrian arab republic" OR tajikistan OR tadjikistan OR tadzhikistan OR tadzhik OR tanzania OR tanganyika OR thailand OR siam OR "timor leste" OR "east timor" OR togo OR "togolese republic" OR tonga OR trinidad OR tobago OR tunisia OR turkey OR turkmenistan OR turkmen OR uganda OR ukraine OR uzbekistan OR uzbek OR vanuatu OR "new hebrides" OR venezuela OR vietnam OR "viet nam" OR "middle east" OR "west bank" OR gaza OR palestine OR yemen OR yugoslavia OR zambia OR zimbabwe OR "northern rhodesia" OR "global south" OR "africa south of the sahara" OR "sub saharan africa" OR "subsaharan africa" OR "africa, central" OR ("central africa" or "africa, northern" OR "north africa" OR "northern africa" OR magreb OR maghrib OR sahara OR "africa, southern" OR "southern africa" OR "africa, eastern" OR "east africa" OR "eastern africa" OR"africa, western" OR "west africa" |

| | |
|---|---|
| | OR "western africa" OR "west indies" OR "indian ocean islands" OR caribbean OR "central america" OR "latin america" OR "south and central america" OR "south america" OR "asia, central" OR "central asia" OR "asia, northern" OR "north asia" OR "northern asia" OR "asia, southeastern" OR "southeastern asia" OR "south eastern asia" OR "southeast asia" OR "south east asia" OR "asia, western" OR "western asia" OR "europe, eastern" OR "east europe" OR "eastern europe" OR "developing country" OR "developing countries" OR "developing nation" OR "developing nations" OR "developing population" OR "developing populations" OR "developing world" OR "less developed country" OR "less developed countries" OR "less developed nation" OR "less developed nations" OR "less developed population" OR "less developed populations" OR "less developed world" OR "lesser developed country" OR "lesser developed countries" OR "lesser developed nation" OR "lesser developed nations" OR "lesser developed population" OR "lesser developed populations" OR "lesser developed world" OR "under developed country" OR "under developed countries" OR "under developed nation" OR "under developed nations" OR "under developed population" OR "under developed populations" OR "under developed world" OR "underdeveloped country" OR "underdeveloped countries" OR "underdeveloped nation" OR "underdeveloped nations" OR "underdeveloped population" OR "underdeveloped populations" OR "underdeveloped world" OR "middle income country" OR "middle income countries" OR "middle income nation" OR "middle income nations" OR "middle income population" OR "middle income populations" OR "low income country" OR "low income countries" OR "low income nation" OR "low income nations" OR "low income population" OR "low income populations" OR "lower income country" OR "lower income countries" OR "lower income nation" OR "lower income nations" OR "lower income population" OR "lower income populations" OR "underserved country" OR "underserved countries" OR "underserved nation" OR "underserved nations" OR "underserved population" OR "underserved populations" OR "underserved world" OR "under served country" OR "under served countries" OR "under served nation" OR "under served nations" OR "under served population" OR "under served populations" OR "under served |

| | |
|---|---|
| | world" OR "deprived country" OR "deprived countries" OR "deprived nation" OR "deprived nations" OR "deprived population" OR "deprived populations" OR "deprived world" OR "poor country" OR "poor countries" OR "poor nation" OR "poor nations" OR "poor population" OR "poor populations" OR "poor world" OR "poorer country" OR "poorer countries" OR "poorer nation" OR "poorer nations" OR "poorer population" OR "poorer populations" OR "poorer world" OR" developing economy" OR" developing economies" OR "less developed economy" OR "less developed economies" OR" lesser developed economy" OR "lesser developed economies" OR "under developed economy" OR "under developed economies" OR "underdeveloped economy" OR "underdeveloped economies" OR "middle income economy" OR "middle income economies" OR "low income economy" OR "low income economies" OR "lower income economy" OR "lower income economies" OR "low gdp" OR "low gnp" OR "low gross domestic" OR "low gross national" OR "lower gdp" OR "lower gnp" OR "lower gross domestic" OR "lower gross national" OR lmic OR lmics OR "third world" OR "lami country" OR "lami countries" OR "transitional country" OR "transitional countries" OR "emerging economies" OR "emerging nation" OR "emerging nations") |
| Social media or internet | “Social media” OR “Social media platform” OR “Social media website” OR “online platform” OR “online site” OR “Online” OR” Virtual “OR “internet” OR “internet use” OR “social network site” OR “technolog*” OR “Digital platform” OR ”WhatsApp” OR “Facebook” OR “Twitter” OR “Instagram” OR “Tik Tok” OR “Telegram” OR “Chatroom” OR “Web” |
